# Genetic Counselor Utilization Across Non-Genetics Departments for Neurodevelopmental Disorders

**DOI:** 10.64898/2026.07.20.26358492

**Authors:** Jordan J. Cole, Julie S. Cohen, Mustafa Sahin, Siddharth Srivastava, Colleen A. Campbell, the Intellectual & Developmental Disabilities Research Centers (IDDRC), Advocating for Access to Genomic Testing Workgroup

## Abstract

**IMPORTANCE:** Most United States children with neurodevelopmental disorders have not received genetic testing aligned with current guidelines. Integration of genetic counselors into non-genetics departments is a potential strategy to improve uptake, but prevalence and details of integrated care models are unknown.

**OBJECTIVE:** To characterize availability, utilization, and perceived need for genetic counselors across non-genetics departments caring for patients with neurodevelopmental disorders

**DESIGN:** Cross-sectional observational department-level survey

**SETTING:** Child neurology, adult neurology, developmental pediatrics, child psychiatry, and adult psychiatry departments at Intellectual and Developmental Disabilities Research Centers

**PARTICIPANTS:** The survey was distributed to 67 departments across 15 institutions. The departmental response rate was 52% (35/67), with at least one response from 87% (13/15) of institutions.

**EXPOSURE:** Presence/absence of dedicated genetic counselor(s), where “dedicated” was defined as hired by the department

**MAIN OUTCOME(S) AND MEASURE(S):** This was a descriptive study only, with no comparative statistical analyses due to the exploratory nature.

**RESULTS:** One third of departments (34%; 12/35) reported having dedicated clinical genetic counselors. Prevalence was highest in child neurology (67%; 8/12), followed by adult neurology (40%; 2/5) and developmental pediatrics (22%; 2/9), with none in child psychiatry (0/7) or adult psychiatry (0/2). In almost all departments with genetic counselors (92%; 11/12), they directly billed for their services, which universally included pre-test counseling/consent and post-test counseling. In departments without genetic counselors, only 39% (9/23) reported providers ordered their own genetic testing. Among all departments, over half (57%) were interested in adding/increasing genetic counseling support, while 26% were unsure and 17% uninterested. Insufficient funding was the most cited barrier; only one department reported insufficient need.

**CONCLUSIONS AND RELEVANCE:** Though currently implemented in only one third of departments, our findings suggest those with dedicated genetic counselors directly pursue genetic testing (without referring to genetics) more than those without genetic counselors. Interest in increasing or adding genetic counseling support was high, and though funding was a reported barrier, feasible funding models were described. In the context of limited medical geneticists and expanding precision therapies, alternate delivery models for neurodevelopmental genetic testing including genetic counselor integration in non-genetics departments may help to scale and sustain uptake.

**Key Points:** *QUESTION:* What is the availability, utilization, and perceived need for genetic counselors in non-genetics departments caring for individuals with neurodevelopmental disorders?

*FINDINGS:* In this cross-sectional study of 35 neurology, psychiatry, and developmental pediatrics departments, one third reported having dedicated genetic counselors for clinical care. Most were interested in increasing or adding genetic counselor support; insufficient funding was the most reported barrier and only one department reported insufficient need.

*MEANING:* Many non-genetics departments caring for individuals with neurodevelopmental disorders continue to rely on the traditional referral model to genetics departments for testing/counseling despite substantial interest and support for integrating genetic counselors.

## Introduction

Neurodevelopmental disorders (NDDs) are childhood-onset conditions that involve a spectrum of differences in typical developmental trajectories in one or more domains (e.g., motor, communication, cognition, etc.), leading to impaired functioning across settings.^1^ Approximately 1 in 6 children in the United States has been diagnosed with an NDD.^2–4^ Epilepsy can also often be conceptualized as an NDD; many individuals meet full criteria and carry NDD diagnoses while many more experience subthreshold or undiagnosed symptoms/disorders, and there is substantial overlap in the genetic background of epilepsy and NDDs.^5^ Among those with epilepsy, autism spectrum disorder, cerebral palsy, and/or intellectual developmental disorder, an underlying genetic disorder is found in approximately 40% with current testing technology.^6–11^ For the purposes of this article, “NDDs” heretofore refers only to these specific conditions unless otherwise stated. Due to the high prevalence of genetic disorders, professional society guidelines from the American Academy of Pediatrics, the American College of Medical Genetics & Genomics, and the National Society of Genetic Counselors/American Epilepsy Society recommend consideration of genetic testing for all individuals with these conditions, and specifically advise first-tier whole exome/genome sequencing for those in whom no obvious acquired etiology or discrete genetic disorder is identified.^6,7,12–14^

Diagnosis of a genetic condition for individuals with NDDs can profoundly impact multiple aspects of medical care, patient/family perspectives, and quality of life.^15–17^ Benefits include access to disease-modifying precision therapies, surveillance for associated health conditions, reproductive counseling, access to advocacy groups, and disease prognostication/planning, among many others. Simply having an answer and ending the “diagnostic odyssey” offers profound meaning for many families.^16^ Often, invasive diagnostic procedures can be avoided.^16^ For those with epilepsy, antiseizure medication choice is informed by an underlying genetic diagnosis in about one third of cases.^18^ In addition, an increasing number of genetic NDDs have FDA-approved disease-specific therapies, with dozens more under active investigation.^19^ Therefore, ensuring prompt access to genetic testing is becoming critical to identify patients who qualify for these novel therapies.

It is essential that patients/families understand the range of potential benefits, so they can be accurately weighed against potential downsides and risks of genetic testing. Pre-test genetic counseling, one of the core roles of genetic counselors (GCs), can aid patients and caregivers in contextualizing these factors to make informed decisions. Beyond direct provision of pre-test counseling, GCs can act as facilitators/educators to frontline clinicians, select the appropriate genetic test, and assist with insurance authorization and other logistics. GCs can also provide comprehensive post-test counseling including accurate result interpretation, result disclosure, diagnosis education, psychosocial support, family planning options, and navigation of cascade testing for family members when applicable.^20^

In the United States, GCs are mid-level healthcare providers who have completed advanced training at Accreditation Council for Genetic Counseling-approved graduate programs, are certified by the American Board of Genetic Counseling, and are licensed in 39 states and Washington DC. Due to their dedicated skills and expertise, there have been calls for and practices demonstrating the success of integrating GCs into clinical care settings beyond the traditional medical genetics specialty departments.^21–24^ Such practices are considered “mainstreaming” models of genetic testing. One of the primary arguments for “mainstreaming” genetic testing is to improve access. Due to shortages of medical geneticists and the growing indications for genetic/genomic testing and precision-based therapies, the traditional model in which genetic testing is pursued only by geneticists is no longer sustainable.^24,25^ There is a clear need for genetic services among individuals with NDDs, as diagnosis rates and indications for genetic testing continue to increase.^4^ The vast majority of individuals with NDDs have not received standard-of-care genetic testing, with many never receiving *any* genetic testing, providing evidence of inadequacy of the traditional genetics service delivery model.^26,27^

In contrast to the limited number of medical geneticists, the number of GCs continues to steadily grow in the U.S.,^28^ offering an opportunity for alternate genetics service delivery. The incorporation of GCs into non-genetics specialties who routinely care for individuals with NDDs, such as neurology, psychiatry, and developmental pediatrics, has begun across many U.S. institutions/health systems. However, little is known regarding the prevalence and economics of such mainstreaming models or the details of GC roles within them.

We sought to address this gap through an inventory of current practice models among the neurology, psychiatry, and developmental pediatrics departments at U.S. institutions designated as Intellectual and Developmental Disabilities Research Centers (IDDRCs).

## Methods

### Regulatory approval/ethics

This study met criteria for IRB exemption at Boston Children’s Hospital (BCH IRB #IRB-P00052583).

### Inventory survey development

A REDCap survey was used to conduct an inventory of genetic counseling service models and needs. The items were developed by members of the IDDRC Workgroup on Advocating for Access to Genomic Testing, which includes 15 total members from nine U.S. institutions, representing child neurology, neurodevelopmental disabilities, child psychiatry, development behavioral pediatrics, medical genetics, and genetic counseling. Most members are currently or previously affiliated with an IDDRC site, and all have clinical and/or research interest in provision of genetic services for children with NDDs. The core team for this project (JJC, SS, JSC, and CAC) developed an initial survey draft. To ensure face- and content-validity, it was revised and finalized (**eFile1**) through an iterative process with feedback from the full workgroup and IDDRC directors.

### Study population

The study population was clinical departments of interest (child neurology, adult neurology, developmental pediatrics, child psychiatry, and adult psychiatry) affiliated with 15 IDDRC sites (**eTable1**). IDDRCs were established in 1963 as centers of excellence for research in intellectual and developmental disabilities, and are funded through the Eunice Kennedy Shriver National Institute for Child Health and Human Development at the National Institutes of Health.^29^

These five clinical departments were chosen due to high prevalence of patients with NDDs. We use the term “department” to represent each specialty *separately*, regardless of whether they are technically part of the same overarching university department. This was to ensure we obtained information at the specialty level, given known differences in patient populations, anticipated differences in GC usage, and the usual presence of distinct clinical leadership. Surveys were completed by department representatives with knowledge of the information being elicited.

### Quantitative survey analysis

Descriptive statistics were completed to reflect frequencies/proportions among all respondents combined and across departments.

## Results

### Response rate

Among the 67 unique departments of interest across the 15 IDDRC sites (**eTable1**), 35 (52.2%) completed surveys. Response rates per specialty were 80.0% (12/15) for child neurology, 60.0% (9/15) for developmental pediatrics, 46.7% (7/15) for child psychiatry, 45.5% (5/11) for adult neurology, and 18.2% (2/11) for adult psychiatry (**Figure 1**). Responses that were duplicates (n=2) or from other departments (n=2) were excluded.

**Figure 1:**
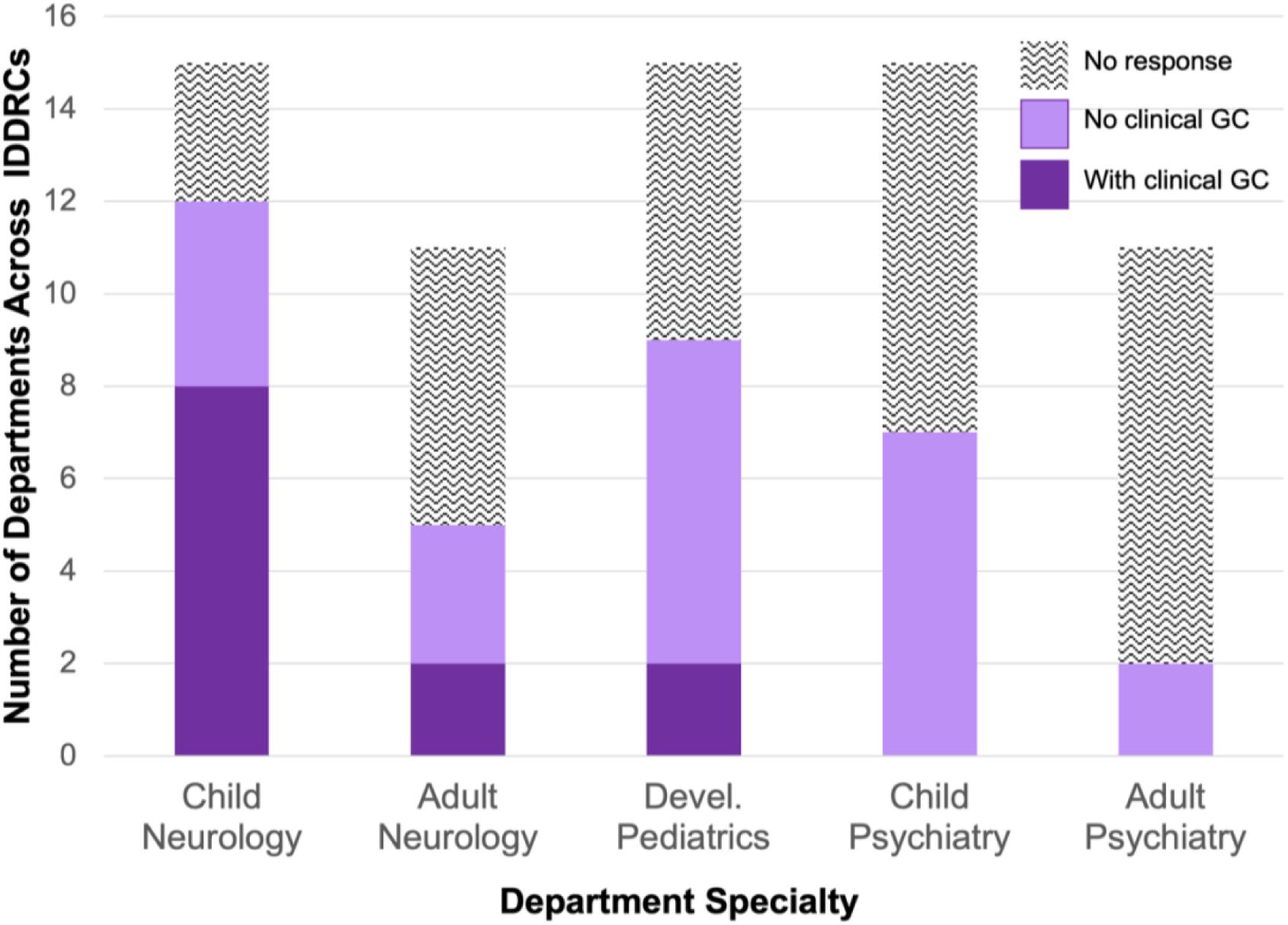
Survey Response Rate & Clinical Genetic Counselor Usage by Department Specialty. *Devel.=Developmental. GC=Genetic Counselor. IDDRCs=Intellectual & Developmental Disabilities Research Centers*.

### Participants

Among the 35 included responses, 13/15 (86.7%) IDDRC sites and 5/5 (100%) clinical departments of interest were represented. **eTable2** contains information on region, specialty, and survey completer roles.

### Clinical genetic counseling workforce

Respondents were instructed to consider GCs or genetic counseling assistants (GCAs) as “dedicated to” or “within” their department only if the GC or GCA was hired by that department. Approximately one third (12/35, 34.3%) reported having dedicated GCs conducting clinical care services (“clinical-GCs”). By specialty, clinical-GCs were present in 8/12 (66.7%) child neurology, 2/5 (40.0%) adult neurology, 2/9 (22.2%) developmental pediatrics departments, and no psychiatry departments **(Figure 1)**. Among departments with clinical-GCs, 5/12 (41.6%) also had clinical-GCAs. One had a clinical-GCA but no clinical-GC.

### Research genetic counseling workforce

A total of 12/35 (34.3%) of departments reported having dedicated GCs conducting or supporting research (“research-GCs”). Two thirds (8/12, 66.7%) with research-GCs also had clinical-GCs. By specialty, research-GCs were present in 3/5 (60.0%) adult neurology, 4/9 (44.4%) developmental pediatrics, 4/12 (33.3%) child neurology, 1/7 (14.3%) child psychiatry, and no adult psychiatry departments. Among departments with research-GCs, 7/12 (58.3%) also had research-GCAs. One had a research-GCA but no research-GC.

### Clinical time, billing practices, and funding mechanisms for clinical-GCs

Among the 12 departments with clinical-GCs, the total number of clinical full-time equivalents (cFTE) per department ranged from 0.25 to 8.0 cFTE (mean=2.6). The estimated average number of patients seen monthly by clinical-GCs per 1.0 cFTE ranged from 10 to 61.5 (mean=43.8; two missing responses).

The vast majority (11/12, 91.7%) of departments with clinical-GCs endorsed multimodal funding models that included direct GC billing plus hospital-, department-, and/or grant-based funding **(Figure 2)**. Only one reported clinical-GCs did *not* bill for clinical services. Conversely, two reported that GC billing was the *only* source of funding for clinical-GC services.

**Figure 2:**
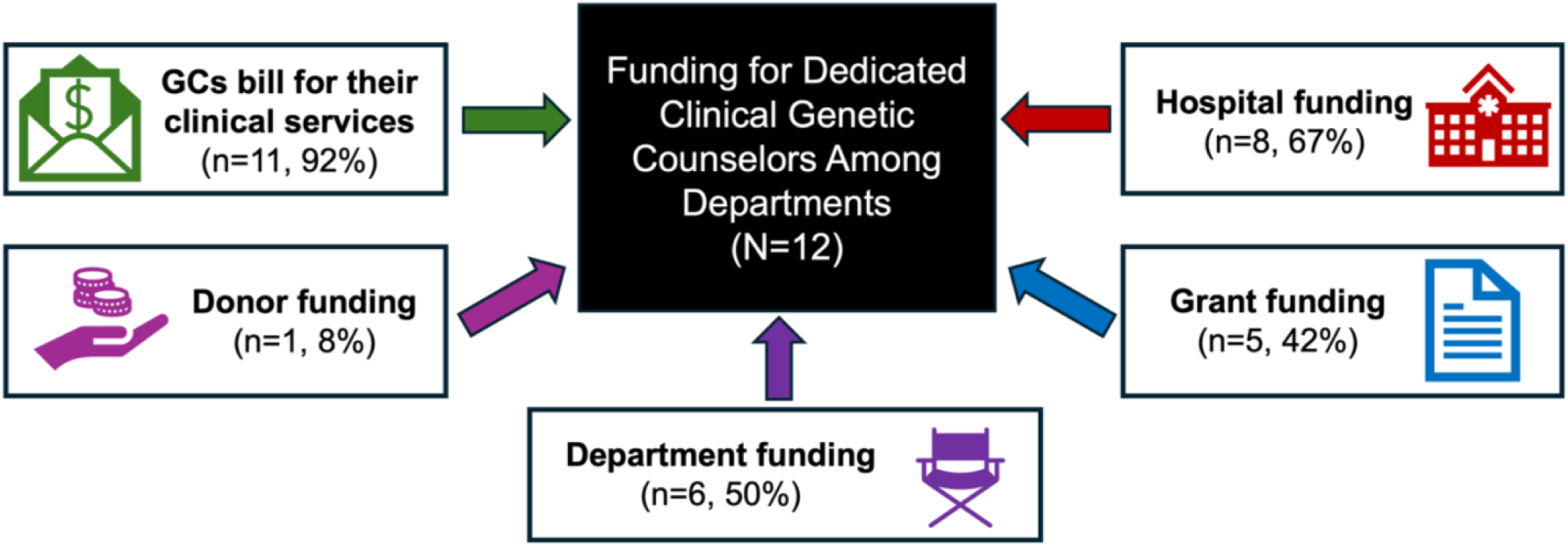
Departments’ Reported Funding Sources for Clinical Genetic Counselors. Departments were asked to select all that applied from a list of possible funding sources and were also given a free response option. Almost all reported that genetic counselor (GC) billing for clinical services accounted for at least partial funding. Most also endorsed additional funding sources, primarily from their department and/or hospital, though some from grants and one from donor(s).

Among the 11 departments utilizing GC billing, eight provided information about Current Procedural Terminology (CPT®) codes **(eTable3)**. All reported using the genetic counseling CPT® code 96040/96041 (before/after 1/1/25). Four free responses indicated use of other CPT® codes in certain scenarios (99211 when insurance excludes 96041 and visit occurs on a subsequent day from the referring physician; 99417 when insurance excludes 96041 and clinical-GC sees the patient with a physician; and G0463 for patients with Medicare/Medicaid). Regarding National Provider Identifier (NPI) numbers used, 8/11 (72.7%) provided this information, while two were unsure and one left it blank. Excluding the unsure/missing responses, 8/8 (100%) departments reported clinical-GCs billed using their own NPI at least sometimes. Half also endorsed physician NPI usage and one quarter hospital NPI usage. One participant specified that the NPI used depended on whether billing was “incident to” [a physician] or “independent”.

### Visit types and roles of clinical-GCs

Among departments with clinical-GCs, all but one provided information on clinical-GC visit types. Most (9/11, 81.8%) reported both in-person and telemedicine visits; two reported only in-person. Regarding clinical-GC roles **(Figure 3)**, all departments indicated provision of pre-test counseling and consenting and post-test counseling. Over half (7/12, 58.3%) specifically endorsed discussion of reproductive risk/options. Fewer than half (5/12, 41.7%) reported facilitating insurance authorizations.

**Figure 3:**
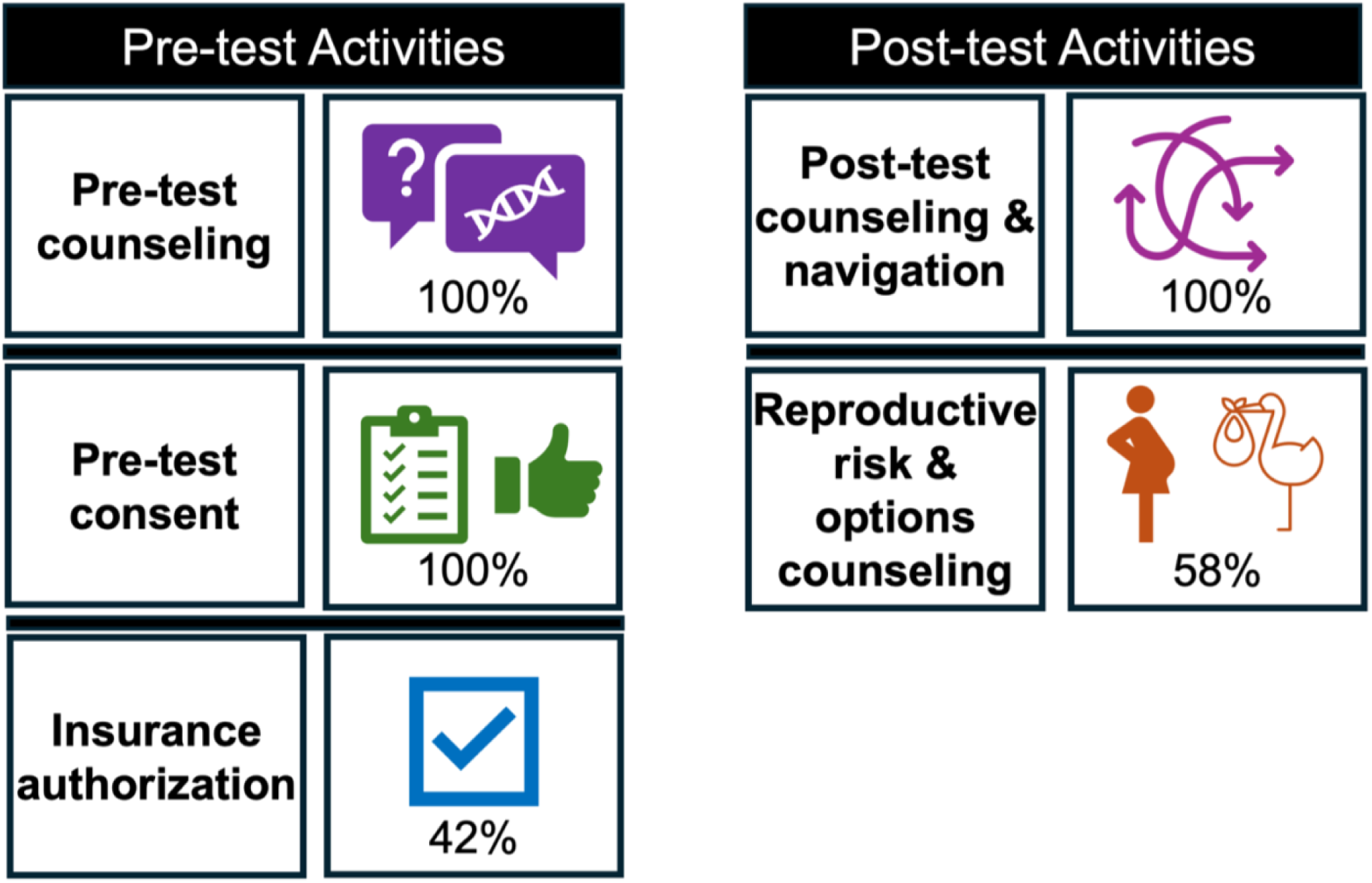
Departments’ Reported Roles of Clinical Genetic Counselors. Respondents could select from a short list of possible roles performed by clinical genetic counselors (GCs) and/or enter free responses. All free responses fell into the above categories but provided more task details, which included result disclosure, facilitating referrals, cascade genetic testing, functional testing, and connecting families with resources related to diagnoses or undiagnosed status.

### Reported barriers among departments without clinical-GCs

Among the 23 departments who reported no clinical-GCs, respondents were asked to indicate whether insufficient need, insufficient funding, and/or another reason prevented them from hiring clinical-GCs. Only one (1/23, 4.3%) endorsed insufficient need, an adult psychiatry department. Approximately half (11/23, 47.8%) reported insufficient funding. Among those who endorsed neither insufficient need nor insufficient funding, most (9/12, 75.0%) provided free responses indicating they referred patients to genetics for genetic services. Two free responses indicated lack of leadership support. One child psychiatry department clarified they *did* have access to specifically-assigned GCs, but they were not technically hired by their department, which was how this study defined “dedicated”.

### Methods of obtaining genetic testing among departments without clinical-GCs

Departments without clinical-GCs were asked how providers obtained genetic testing. Most (18/23, 78.3%) indicated referring patients to the genetics department; this was the *only* indicated method for 7/23 (30.4%). Another method was referral to GCs outside the department (9/23, 39.1%). Only nine (39.1%) reported their clinicians obtained genetic testing on their own, without formal genetic counseling by a certified GC. Some provided details that this was done only for particular test types with which providers were comfortable counseling and interpreting (e.g., chromosomal microarray and Fragile X testing in a developmental behavioral pediatrics department, Huntington’s disease testing in an adult psychiatry department).

### Interest in increasing or adding clinical-GCs

Among the 12 departments with clinical-GCs, most (9/12, 75%) indicated interest in increasing GC’s’ cFTE, while two were unsure (16.7%), and one was uninterested (8.3%). Among the 23 departments without clinical-GCs, nearly half (11/23, 47.8%) indicated interest in adding GCs, while the rest were unsure (7/23, 30.4%) or uninterested (5/23, 21.7%). **Figure 4** demonstrates clinical-GC utilization and interest by specialty.

**Figure 4:**
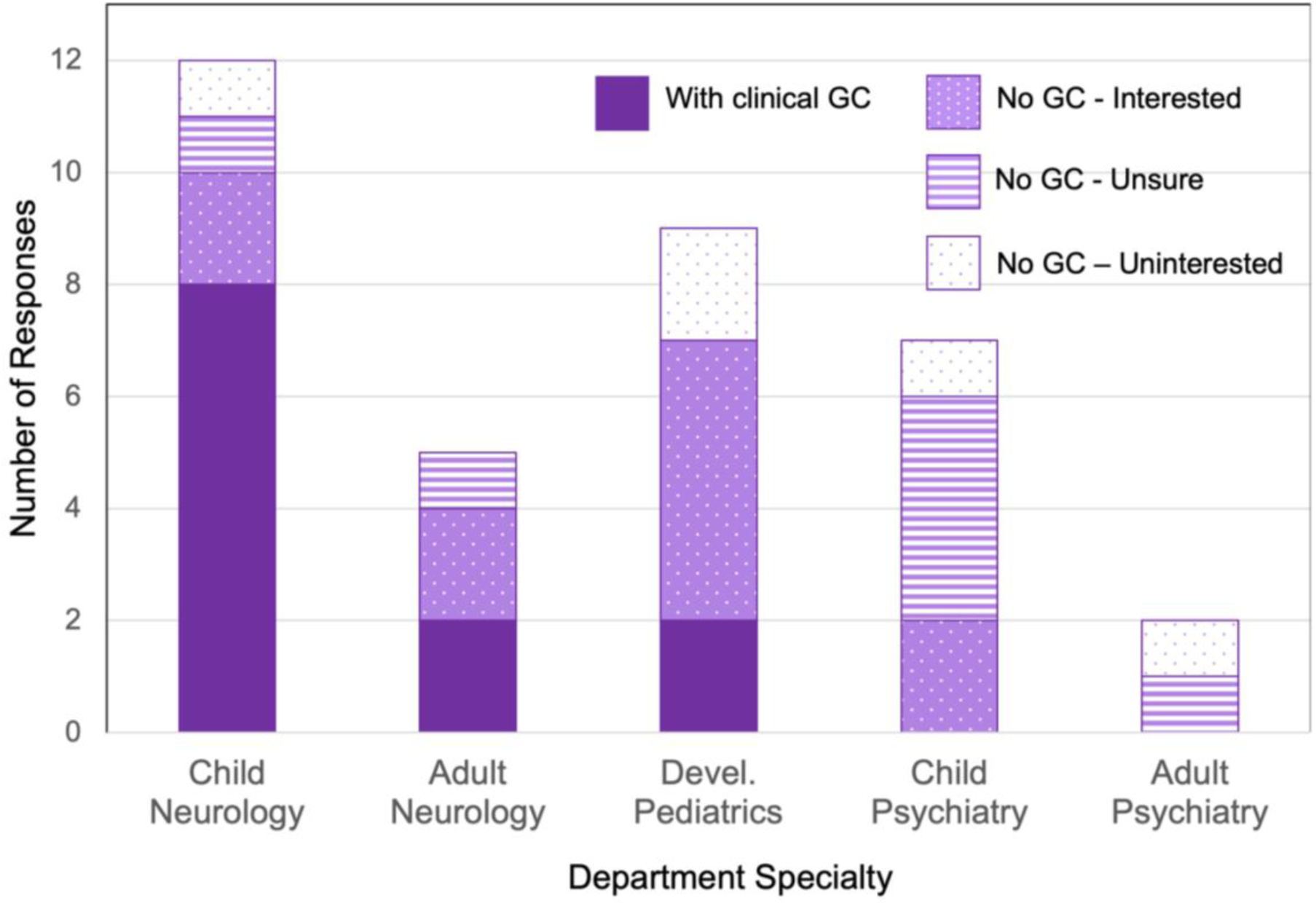
Clinical Genetic Counselor Utilization & Interest by Department Specialty. *Devel.=Developmental. GC=Genetic Counselor*.

## Discussion

As the recommendations for genetic testing in individuals with NDDs expand and potential benefits of genetic diagnosis increase, it is essential that alternate genetic service delivery models are explored to ensure accessibility. Prior publications have described the concept of “mainstreaming” genetic testing, in which all or part of the clinical genetic testing process is completed outside the genetics department,^24^ and studies have reported non-genetics clinicians’ NDD genetic testing practices.^30,31^ However, none has explicitly focused on delineating the prevalence, roles, and billing practices of dedicated GCs supporting/facilitating NDD genetic testing in non-genetics departments. Our study sought to achieve this through a survey of neurology, psychiatry, and developmental pediatrics departments at IDDRC sites.

Overall, we found that approximately 1/3 of departments currently employ clinical-GCs. The proportion varied substantially by specialty, with 66.7% of child neurology, 40% of adult neurology, and 22.2% of developmental pediatrics departments endorsing use of dedicated clinical-GCs, while none of the psychiatry departments did. Higher rates in neurology may relate to higher prevalence of epilepsy, movement disorders, or neuromuscular disorders, or it may reflect presence of clinical-GCs in clinics dedicated to specific neurogenetic conditions such as Rett syndrome or neurofibromatosis. The relatively low clinical-GC prevalence in developmental pediatrics may relate to developmental pediatrics workforce shortages^32^ – resources may be directed towards hiring clinicians or neuropsychologists rather than clinical-GCs, though this is speculative. The absence of clinical-GCs within psychiatry may reflect a primary focus on management of mood/behavior rather than etiologic evaluation, and/or could relate to the polygenic nature of most psychiatric conditions.^33^

The variability in clinical-GCs by specialty may also relate to differences in genetic education during residency/fellowship and/or differences in genetic content among academic resources and society guidelines. Though trainees and licensed clinicians across specialties perceive need for increased genetics education,^34–37^ genetic didactics/practical training is particularly sparse in psychiatry.^38^ The Diagnostic and Statistical Manual of Mental Disorders, 5^th^ Edition, the standard diagnostic resource used by psychiatrists, does not include specification of genetic etiologies in neurodevelopmental disorders criteria.^39^ In contrast, the International League Against Epilepsy papers on epilepsy syndromes explicitly discuss genetic etiologies and the importance of genetic testing.^40,41^ As to academic society guidelines, there are significant discrepancies across specialties regarding indications and first-tier testing types.^42^

All departments with clinical-GCs reported roles involving pre-test and post-test genetic counseling, implying 100% of these departments had at least some providers ordering genetic testing without the direct involvement of medical geneticists. In contrast, among departments *without* clinical-GCs, only 39% reported their providers order their own genetic testing, and several specified that providers only ordered particular tests with which they were comfortable interpreting and providing counseling. Prior studies have also shown an association between genetic test ordering and access to GCs among non-geneticists.^37,43,44^ This may relate to GCs reducing barriers by handling testing logistics and/or the frequent insurance requirement for formal genetic counseling for whole exome/genome sequencing.^45^ It may also increase provider willingness to pursue genetic testing due to the added layer of expertise GCs provide regarding test selection, variant interpretation, and psychosocial counseling.^46^ In a national survey of U.S. neurogeneticists, hiring GCs/GCAs was the highest recommended step to developing an effective neurogenetics program.^47^ In our study, most departments with clinical-GCs endorsed desire to increase clinical-GC support, suggesting perceived benefits and continued need for these services.

Interestingly, among departments without clinical-GCs, many did not report either insufficient funding or insufficient need as reasons, instead simply citing use of the traditional genetic care model involving referral to genetics. We did not assess whether respondents were satisfied with the traditional model, nor what the wait times and rates of completed genetics referrals were. Prior studies have shown that many patients never follow through to completing visits after genetics referrals, suggesting that genetic testing may be more accessible when available through established providers.^48^ Many did endorse interest in adding dedicated clinical-GCs, suggesting they are not fully satisfied with the traditional model. It is possible that insufficient funding was not reported as a barrier if the traditional model was accepted as the status quo and the logistics of adding clinical-GCs had not been explored.

We found that the vast majority of departments with clinical-GCs (all but one) utilize GC billing to fund the positions. This is an important finding due to known issues with reimbursement for GC services, in part related to lack of recognition of GCs as medical providers by Medicare and Medicaid.^49^ It suggests that despite barriers surrounding reimbursement, GC billing does provide at least partial financial benefit. There are ongoing advocacy efforts at the state and national levels to formally recognize GCs as medical providers, which is anticipated to improve overall reimbursement and may mitigate funding barriers.^50,51^

There are several limitations of this study. To promote feasibility and survey completion, the survey items included a limited number of pre-written options, requiring respondents to use free-text response to add additional/other information. Thus, the full scope of genetic testing/counseling practices and roles may not be represented. We also relied on sites to self-identify the most appropriate person to complete the survey, and it is possible there are inaccuracies or inconsistencies. The overall response rate was 52.2%, and while responding departments were spread across 13/15 IDDRC sites, two sites had no responses, and it is not known whether respondents differed from non-respondents in important ways. Furthermore, IDDRC sites may not be representative of the larger U.S. health care system with regards to specialty services for NDDs. This was an intentional population chosen for study due to the desire to characterize practices at sites actively pursuing advancements in NDD research. Future directions may include follow-up interviews with respondents of the survey reported in this article, and/or expansion to non-IDDRC sites.

## Conclusion

This study provides details of health service models involving dedicated GCs in non-genetics specialty departments. Though currently implemented in only 1/3 of surveyed departments, our findings suggest those with dedicated genetic counselors directly pursue genetic testing (without referring to genetics) more than those without genetic counselors. Most departments were interested in adding or increasing genetic counseling support, and among those currently using GCs, the vast majority reported successful GC billing for clinical services as a component of funding. We found strong support for utility and feasibility of GC embedment within non-genetics departments that provide care for individuals with NDDs. This will be increasingly important as the potential benefits of NDD genetic testing continue to increase due to continued discovery of disease genes, improved prognostication with natural history studies, increased availability of reproductive options, and development of precision therapies.

## Data Availability

The data that support the findings of this study are available from the corresponding author upon reasonable request.

## Acknowledgements

Each of the authors (JJC, JSC, MS, SS, and CAC) met authorship criteria according to the journal guidelines. JJC contributed to the study conception/design, data acquisition/analysis, interpretation, and drafting the manuscript. MS contributed to data acquisition and interpretation and critically reviewing the manuscript. JSC, SS, and CAC contributed to the study conception/design, data acquisition/analysis, interpretation, and critically reviewing the manuscript. All authors approve the final version for publication and agree to be accountable for all aspects of the work.

Additional members of the Intellectual & Developmental Disabilities Research Centers (IDDRC) Advocating for Access to Genomic Testing Workgroup who served as nonauthor collaborators for this project include (listed alphabetically): Julia S. Anixt, MD (Cincinnati Children’s Hospital and University of Cincinnati Department of Pediatrics, Division of Developmental & Behavioral Pediatrics); Maya Chopra, MBBS (Boston Children’s Hospital and Harvard University Department of Neurology, Division of Genetics & Genomics); Matthew Deardorff, MD, PhD (Children’s Hospital Los Angeles and University of Southern California Departments of Pediatrics and Clinical Pathology); Christina Gurnett, MD, PhD (Children’s Hospital Philadelphia and University of Pennsylvania Department of Pediatrics, Division of Neurology); Shafali Jeste, MD (UCLA Mattel Children’s Hospital and University of California Los Angeles Departments of Pediatrics and Neurology); Ernest Pedapati, MD, MS (Cincinnati Children’s Hospital and University of Cincinnati Department of Psychiatry, Division of Child & Adolescent Psychiatry); and Beth Rosen Sheidley, MS, CGC (Boston Children’s Hospital Department of Neurology). These individuals contributed to the study conceptualization and to the iterative refinement of the survey items.

## Funding/Support

JJC receives research support through the National Institute of Neurological Disorders & Stroke (NINDS) at the National Institutes of Health (NIH) (5K12NS098482). JSC receives research support through the National Institute of Child Health and Human Development (NICHD) at NIH (P50HD103538). MS is funded by Boston Children’s Hospital Intellectual and Developmental Disabilities Research Center (P50HD105351). SS receives research support through the NICHD at the NIH (1R01HD121615).

The NIH was not involved in study design, data collection/analysis, interpretation of the data, or any aspects of manuscript preparation/submission.

## Data Access, Responsibility, and Analysis

JJC, SS, and CAC had full access to all the data in the study and take responsibility for the integrity of the data and the accuracy of the data analysis.

## Data Sharing Statement

The data that support the findings of this study are available from the corresponding author upon reasonable request. The data are not publicly available for privacy reasons.

