## Supplemental Materials for "Genetic Counselor Utilization Across Non-Genetics Departments for Neurodevelopmental Disorders"

**eFile1: Final Inventory Survey**…………………………………………………………………………………………..…page 2-5

**eTable1: List of IDDRC Sites and the Clinical Departments of Interest Available at Each**…………………………………...page 6

**eTable2: Location, Specialty, & Survey Completer Roles Among Participating Departments**……………………………….page 7

**eTable3: Current Procedural Terminology (CPT®) Codes Used by Clinical Genetic Counselors**……………………………page 8

**Abbreviations:**

Cont.: Continued

CPT: Current Procedural Terminology

E/M: Evaluation/Management

FTE: Full time equivalent

GC: Genetic counselor

IDDRC: Intellectual and Developmental Disabilities Research Center(s)

NPI: National provider identifier

### eFile1: Final Inventory Survey

**SURVEY INTRODUCTION**

Thank you for completing this inventory survey. It should take < 5 minutes to complete.

Purpose: To survey the landscape of genetic counseling utilization within IDDRC sites for patients with neurodevelopmental disorders (developmental delay, intellectual disability, autism spectrum disorder, cerebral palsy, and epilepsy).

Instructions: Please complete a separate inventory survey for each department listed below. It is ok to forward this inventory to other people within your IDDRC for completion.

- Adult neurology
- Child neurology
- Adult psychiatry
- Child psychiatry
- Developmental behavior pediatrics

Incentive: You will later receive a compiled report of the results (pertaining to your individual site and all the sites in aggregate) that can be incorporated into annual IDDRC reports.

PLEASE REVIEW BEFORE BEGINNING: Important Definitions of Terms Used in This Survey

- Department = Consider each specialty listed above as a separate department for the purposes of this study, regardless of whether it is technically a division or section of a larger department
- Dedicated = Hired by the department about which you are completing this inventory. Do NOT include genetic counselors who work within your department but are hired by another department.
- Genetic counselor assistant = Person who helps with organizational, administrative, research, or insurance-related tasks, often under the direct supervision of a genetic counselor.

**SURVEY SECTION A: INFO ON IDDRC SITE AND PERSON COMPLETING SURVEY**

| Item # | Item Text | Response Options |
|---|---|---|
| A1 | Your name | *[free text]* |
| A2 | Your email | *[free text]* |
| A3i | Your role in IDDRC | *[select ONE]*<br>• Director<br>• Other |
| A3ii | *[shown only if response to A3i = Other]*<br>Please specify | *[free text]* |
| A4 | About which IDDRC site are you providing information? | *[select ONE]*<br>• Albert Einstein College of Medicine<br>• Baylor College of Medicine<br>• Boston Children's Hospital<br>• Children's Hospital of Philadelphia<br>• Children's National Medical Center<br>• Kennedy Krieger Institute<br>• University of California Davis<br>• University of California Los Angeles<br>• University of Iowa<br>• University of North Carolina<br>• University of Rochester<br>• University of Washington<br>• University of Wisconsin<br>• Vanderbilt University<br>• Washington University |
| | *Continued on next page* | |

| Item # cont. | Item Text cont. | Response Options cont. |
|---|---|---|
| A5i | About which department are you completing the survey?<br><br><u>Reminder</u>: Please complete a separate instance of the survey for each department. | *[select ONE]*<br>• Adult neurology<br>• Child neurology<br>• Adult psychiatry<br>• Child psychiatry<br>• Developmental behavioral pediatrics<br>• Other |
| A5ii | *[shown only if response to A5i = Other]*<br>Please specify | *[free text]* |

**<u>SURVEY SECTION B</u>: YES/NO – DEDICATED GENETIC COUNSELORS FOR CLINICAL CARE**

| Item # | Item Text | Response Options |
|---|---|---|
| B1 | Does this department utilize dedicated genetic counselors for clinical care? | *[select ONE]*<br>• Yes<br>• No |
| ***BRANCH POINT*** *– Those who select "Yes" on B1 go to Section C. Those who select "No" go to Section D.* | | |

**<u>SURVEY SECTION C</u>: DETAILS OF GENETIC COUNSELOR UTILIIZATION FOR CLINICAL CARE**

*This section is only shown/given to those who selected "Yes" on item B1 (indicating the department utilizes dedicated genetic counselors for clinical care).*

| Item # | Item Text | Response Options |
|---|---|---|
| C1 | In this department, what is the total number of dedicated Full Time Equivalent (FTE) genetic counselors for clinical care?<br><br>Please provide your response in the format 1.0, 2.0, 3.25, 12.5, etc., where 1.0 FTE = standard full-time work in this department. | *[free text]* |
| C2 | In this department, what is the approximate total number of patients seen by all dedicated genetic counselors in a one-month period? | *[free text]* |
| C3 | Would this department like to increase the number of dedicated Full Time Equivalent (FTE) genetic counselors for clinical care? | *[select ONE]*<br>• Yes<br>• No<br>• Not sure |
| C4i | In this department, do dedicated genetic counselors bill for any of their clinical services? | *[select ONE]*<br>• Yes<br>• No |
| C4iia | *[shown only if response to C4i = Yes]*<br>In this department, how do dedicated genetic counselors bill for their clinical care services? | *[select ALL that apply]*<br>• Genetic counselor CPT code 96040 (prior to January 1, 2025) or 96041 (after January 1, 2025)<br>• Not sure/Don't know<br>• Other |
| C4iib | *[shown only if "Other" was selected on C4iia]*<br>Please specify | *[free text]* |
| C4iiia | *[shown only if response to C4i = Yes]*<br>In this department, when dedicated genetic counselors bill for their clinical care services, whose NPI(s) do they use? | *[select ALL that apply]*<br>• Genetic counselor's personal NPI<br>• Physician's NPI<br>• Hospital NPI<br>• Not sure/Don't know<br>• Other |
| C4iiib | *[shown only if "Other" was selected on C4iiia]*<br>Please specify | *[free text]* |
| | *Continued on next page* | |

| Item # cont. | Item Text cont. | Response Options cont. |
|---|---|---|
| C5i | In this department, besides any funding through direct billing for their clinical services, are there other funding sources for dedicated genetic counselors' clinical effort? | *[select ALL that apply]*<br>• Hospital funding<br>• Department funding<br>• Grant funding<br>• Donor funding<br>• None<br>• Other |
| C5ii | *[shown only if "Other" was selected on C5i]*<br>Please specify | *[free text]* |
| C6i | In this department, what are the visit types of dedicated genetic counselors (providing clinical care) who see patients? This question pertains only to GC-only visits. | *[select ALL that apply]*<br>• In-person<br>• Telehealth<br>• Other |
| C6ii | *[shown only if "Other" was selected on C6i]*<br>Please specify | *[free text]* |
| C7i | In this department, what clinical care roles do dedicated genetic counselors play? | *[select ALL that apply]*<br>• Pre-test counseling<br>• Pre-test consenting<br>• Facilitation of insurance preauthorization for genetic testing<br>• Post-test counseling<br>• Reproductive risk and options counseling<br>• Other |
| C7ii | *[shown only if "Other" was selected on C7i]*<br>Please specify | *[free text]* |
| C8 | In this department, do dedicated genetic counselors provide clinical care in conjunction with a physician/advanced practice provider (paired simultaneous evaluation) or as an independent provider? | *[select ONE]*<br>• In conjunction with a physician/advanced practice provider<br>• As an independent provider<br>• Both |

**<u>SURVEY SECTION D</u>: PRACTICES & PERSPECTIVES OF DEPARTMENTS WITHOUT DEDICATED GENETIC COUNSELORS FOR CLINICAL CARE**

*This section is only shown/given to those who selected "No" on item B1 (indicating the department does NOT utilize dedicated genetic counselors for clinical care).*

| Item # | Item Text | Response Options |
|---|---|---|
| D1i | What are the reasons why this department does not utilize dedicated genetic counselors for clinical care? | *[select ALL that apply]*<br>• Insufficient need<br>• Insufficient funding<br>• Other |
| D1ii | *[shown only if "Other" was selected on D1i]*<br>Please specify | *[free text]* |
| D2i | Since this department does NOT utilize dedicated genetic counselors for clinical care, how do providers within the department consider genetic testing for their patients? | *[select ALL that apply]*<br>• Referral to clinical genetics<br>• Referral to genetic counselors outside of the department<br>• On their own, without formal genetic counseling<br>• Other |
| D2ii | *[shown only if "Other" was selected on D2i]*<br>Please specify | *[free text]* |
| | *Continued on next page* | |

| Item # cont. | Item Text cont. | Response Options cont. |
|---|---|---|
| D3 | Though this department does NOT currently utilize dedicated genetic counselors for clinical care, does this department have an interest in doing so? | *[select ONE]*<br>• Yes<br>• No<br>• Not sure |

**<u>SURVEY SECTION E</u>: RESEARCH GENETIC COUNSELORS & GENETIC COUNSELOR ASSISTANTS**

*This section is shown/given to all survey respondents regardless of any previous response selections.*

| Item # | Item Text | Response Options |
|---|---|---|
| E1 | Does this department utilize dedicated genetic counselors to assist with or lead research studies? | *[select ONE]*<br>• Yes<br>• No |
| E2 | Does this department utilize dedicated genetic counselor assistants for clinical care? | *[select ONE]*<br>• Yes<br>• No |
| E3 | Does this department utilize dedicated genetic counselor assistants to assist with research studies? | *[select ONE]*<br>• Yes<br>• No |

**<u>SURVEY SECTION F</u>: FINAL COMMENTS/INFORMATION (OPTIONAL)**

*This section is shown/given to all survey respondents regardless of any previous response selections.*

| Item # | Item Text | Response Options |
|---|---|---|
| F1 | If you would like, please provide any additional information about access to dedicated genetic counselors and genetic testing within this department. | *[free text]* |

| eTable1: List of IDDRC Sites and the Clinical Departments of Interest Available at Each | | | | | | |
|---|---|---|---|---|---|---|
| Site (Alphabetical order) | | Child Neurology | Child Psychiatry | Developmental Pediatrics | Adult Neurology | Adult Psychiatry |
| 1 | Albert Einstein University | Yes | Yes | Yes | Yes | Yes |
| 2 | Baylor University | Yes | Yes | Yes | Yes | Yes |
| 3 | Boston Children's Hospital | Yes | Yes | Yes | No | No |
| 4 | Children's Hospital of Philadelphia | Yes | Yes | Yes | No | No |
| 5 | Children's National Hospital | Yes | Yes | Yes | No | No |
| 6 | Kennedy Krieger Institute | Yes | Yes | Yes | No | No |
| 7 | University of California Davis | Yes | Yes | Yes | Yes | Yes |
| 8 | University of California Los Angeles | Yes | Yes | Yes | Yes | Yes |
| 9 | University of Iowa | Yes | Yes | Yes | Yes | Yes |
| 10 | University of North Carolina | Yes | Yes | Yes | Yes | Yes |
| 11 | University of Rochester | Yes | Yes | Yes | Yes | Yes |
| 12 | University of Washington | Yes | Yes | Yes | Yes | Yes |
| 13 | University of Wisconsin | Yes | Yes | Yes | Yes | Yes |
| 14 | Vanderbilt University | Yes | Yes | Yes | Yes | Yes |
| 15 | Washington University in St. Louis | Yes | Yes | Yes | Yes | Yes |

| eTable2: Location, Specialty, & Survey Completer Roles Among Participating Departments | | |
|---|---|---|
| **Descriptors of Survey Responses** | | **Number of Departments (N=35)** |
| **Regional Location of IDDRC Site**[A] | Northeast | 7 (20%) |
| | Midwest | 9 (26%) |
| | South | 11 (31%) |
| | West | 8 (23%) |
| **Department's Clinical Specialty** | Adult Neurology | 5 (14%) |
| | Child Neurology | 12 (34%) |
| | Adult Psychiatry | 2 (6%) |
| | Child Psychiatry | 7 (20%) |
| | Developmental Pediatrics | 9 (26%) |
| **Clinical Role of Survey Completer**[B] | Physician | 23 (66%) |
| | Genetic counselor | 10 (29%) |
| | Other faculty member | 2 (6%) |
| **Leadership Role of Survey Completer**[B,C] | +Leadership position | 27 (77%) |
| | Not in leadership position | 8 (23%) |

*[A] Regions are as defined by the United States Census Bureau.*
*[B] Survey completer = Person who filled out the survey on behalf of the department. Twenty-nine unique individuals completed survey responses; three completed the survey on behalf of multiple departments. Numbers add to 35 to reflect all department responses.*
*[C] Leadership positions defined as IDDRC director, co-director, core leader, or equivalent; institutional or organizational leader of a department, division, section, or equivalent; clinic director*

| eTable3: Current Procedural Terminology (CPT®) Codes Used by Clinical Genetic Counselors | | | |
|---|---|---|---|
| **CPT® Code** | **Name** | **Description** | **Reported Usage** |
| 96040 | Medical genetics and genetic counseling services | Can be used to bill for each 30 minutes of face-to-face time spent with a patient or family to review family history, interpret genetic tests, and provide psychosocial counseling | Used prior to 1/1/2025, when this code was replaced by 96041 |
| 96041 | Medical genetics and genetic counseling services, total time | Can be used to bill for the total time spent on a patient's care on the date of the encounter (including chart preparation, test coordination, and face-to-face activities) | Used since 1/1/2025, when this code replaced 96040 |
| 99211 | Office or other outpatient visit for the evaluation and management (E/M) of an established patient | The only E/M code that does not require face-to-face presence of a physician. For established patients, other provider can bill for brief/straightforward ("Level 1") services. Often used for patient education or "nurse visits" | Used for patients whose insurance excludes 96041 and GC encounter occurs on a subsequent day from that of the referring provider |
| 99417 | Prolonged office or other outpatient services beyond the total time of the primary procedure | Can be used by physician/advanced practice provider to bill for each 15 minutes of time spent beyond the highest level of time-based-billing service (i.e., 99205 for new patients and 99215 for established patients). Includes both face-to-face and non-face-to-face activities completed on day of encounter | Used for patients whose insurance excludes 96041 and GC sees patient together with a physician/advanced practice provider |
| G0463[A] | Hospital outpatient clinic visit for assessment and management of a patient | Can be used specifically by hospital-owned outpatient clinics to bill for the facility fee associated with a patient's clinic visit | Used for patients with Medicare or Medicaid insurance |

*[A] G0563 is technically not a CPT® code but rather a Healthcare Common Procedure Coding System (HCPCS) code. We have included it here as it is also a billing code.*
